# COVID-19 is not an Independent Cause of Death

**DOI:** 10.1101/2022.06.01.22275878

**Authors:** Marcia C Castro, Susie Gurzenda, Cassio M Turra, Sun Kim, Theresa Andrasfay, Noreen Goldman

**Affiliations:** Department of Global Health and Population, Harvard TH Chan School of Public Health, Boston, MA 02115, USA; Demography Department, Cedeplar, Universidade Federal de Minas Gerais, Belo Horizonte, MG 31270-901, Brazil; Leonard Davis School of Gerontology, University of Southern California, Los Angeles, CA 90089, USA; Office of Population Research and Princeton School of Public and International Affairs, Princeton University, Princeton, NJ 08544, USA

**Keywords:** Competing risks, Life expectancy at birth, COVID-19 mortality

## Abstract

The COVID-19 pandemic has had overwhelming global impacts with deleterious social, economic, and health consequences. To assess the COVID-19 death toll researchers have estimated declines in 2020 life expectancy at birth. Because data are often available only for COVID-19 deaths, the risks of dying from COVID-19 are assumed to be independent of those from other causes. We explore the soundness of this assumption based on data from the US and Brazil, the countries with the largest number of reported COVID-19 deaths. We use three methods. One estimates the difference between 2019 and 2020 life tables and therefore does not require the assumption of independence. The other two assume independence to simulate scenarios in which COVID-19 mortality is added to 2019 death rates or is eliminated from 2020 rates. Our results reveal that COVID-19 is not independent of other causes of death. The assumption of independence can lead to either an overestimate (Brazil) or an underestimate (US) of the decline in *e*_*0*_, depending on how the number of other reported causes of death changed in 2020.

## Introduction

Over 6.2 million deaths were attributed to COVID-19 worldwide by April 2022, a number that is likely underestimated. The two countries with the largest number of reported COVID-19 deaths are Brazil and the US, with more than 666 thousand and a million deaths reported respectively as of May 2022. The impact of COVID-19 on life expectancy at birth (*e*_0_) in these countries has received widespread attention from the media: a decline of 1.3-years in Brazil and 1.8-years in the US for 2020 (Castro et al. 2021; Murphy et al. 2021). Declines are even larger in 2021 (Andrasfay and Goldman 2022; Castro et al. 2021).

To examine the impact of COVID-19, researchers have used numbers of deaths and population by age to calculate declines in *e*_0_ (the metric reported here) during a specified period of the pandemic (e.g., 2020 relative to 2019). Life expectancy has numerous advantages over other mortality measures, including its interpretability, comparability over time and place, and the fact that, unlike the death rate, it is unaffected by the age distribution. Estimates of *e*_0_ during the pandemic for both the US and Brazil have varied across studies primarily because the National Center for Health Statistics (NCHS) in the US and the Ministry of Health (MoH) in Brazil have periodically released updated and corrected information. However, as we demonstrate below, another important source of variation stems from differences in the methods used to calculate *e*_0_, often driven by limited data availability.

Government agencies in the US and Brazil generally release mortality data after a substantial lag to ensure completeness and quality. However, during the pandemic, both the NCHS and MoH released information on COVID-19 – but not other – deaths shortly after receipt, permitting researchers to calculate the impact of the pandemic on *e*_0_ without data from all death records. From mid-2020 until November 2021, preliminary data for all causes of death (CoD) started to be released every two weeks in Brazil. Estimates of the impact of COVID-19 on *e*_0_ and years of life lost (YLL) based on these limited data required the assumption that the risks of dying from COVID-19 were independent of the risks of dying from other causes (the presence of COVID-19 did not alter the risks of dying from any other cause) (Chan, Cheng and Martin 2021). Although unrealistic, this assumption pervades the literature – both before the pandemic and currently – primarily because there has been no direct way to assess dependence among causes. Previous applications of the assumption of independence among CoD typically involved determining the consequences of the hypothetical elimination or deletion of a long-standing cause, frequently a chronic disease such as cancer (Beltrán-Sánchez, Preston and Canudas-Romo 2008; Ho 2013; Yashin, Manton and Stallard 1986). In contrast, the sudden arrival of COVID-19 provides a natural experiment that permits a direct assessment of the plausibility of the independence assumption, in this case between COVID-19 and other CoD, in the context of adding rather than eliminating a cause. We explore the consequences of this assumption using data for the US and Brazil.

## Methods

Deaths from all causes by month, age, sex, and CoD for Brazil were extracted from the Mortality Information System, MoH, for 2018-2020, and population projections by age and sex from the Brazilian Institute of Geography and Statistics (Castro et al. 2021). For the US, corresponding data were obtained from CDC Wonder (Centers for Disease Control and Prevention and National Center for Health Statistics 2021).

The methods used to calculate the decline in *e*_0_ are described elsewhere (Castro et al. 2021). Briefly, we used three approaches. First, we constructed period life tables (LT) for 2019 and 2020 that considered all CoD by sex and age group and calculated the difference between *e*_0_ in 2019 and 2020 (LT19-LT20). This method does not make assumptions about independence among causes or depend on the accurate classification of causes; however, the estimate would include reductions and increases in numbers of deaths not related to the COVID-19 pandemic. The other approaches simulate hypothetical scenarios in which COVID-19 mortality is added to 2019 death rates (DT19) or is eliminated from 2020 rates (DT20); both assume independence between the risks of dying from COVID-19 and those from other causes (Chiang 1968). We estimated reductions in *e*_0_ due to COVID-19 from the difference between the 2020 period LT and the DT20 estimates, and between the 2019 period LT and the DT19 estimates for 2020.

We performed all analyses in R v.4.0.0 (R core team, 2020). We created data visualizations in R and Adobe Illustrator CS6.

## Results

Based on LT19-LT20, the US lost 1.56 years compared to 1.41 in Brazil. Estimates of the change in *e*_0_ derived from DT19 and DT20 under the assumption of independence are larger than LT19-LT20 for Brazil but smaller than LT19-LT20 in the US (**Table 1**).

**Table 1.**
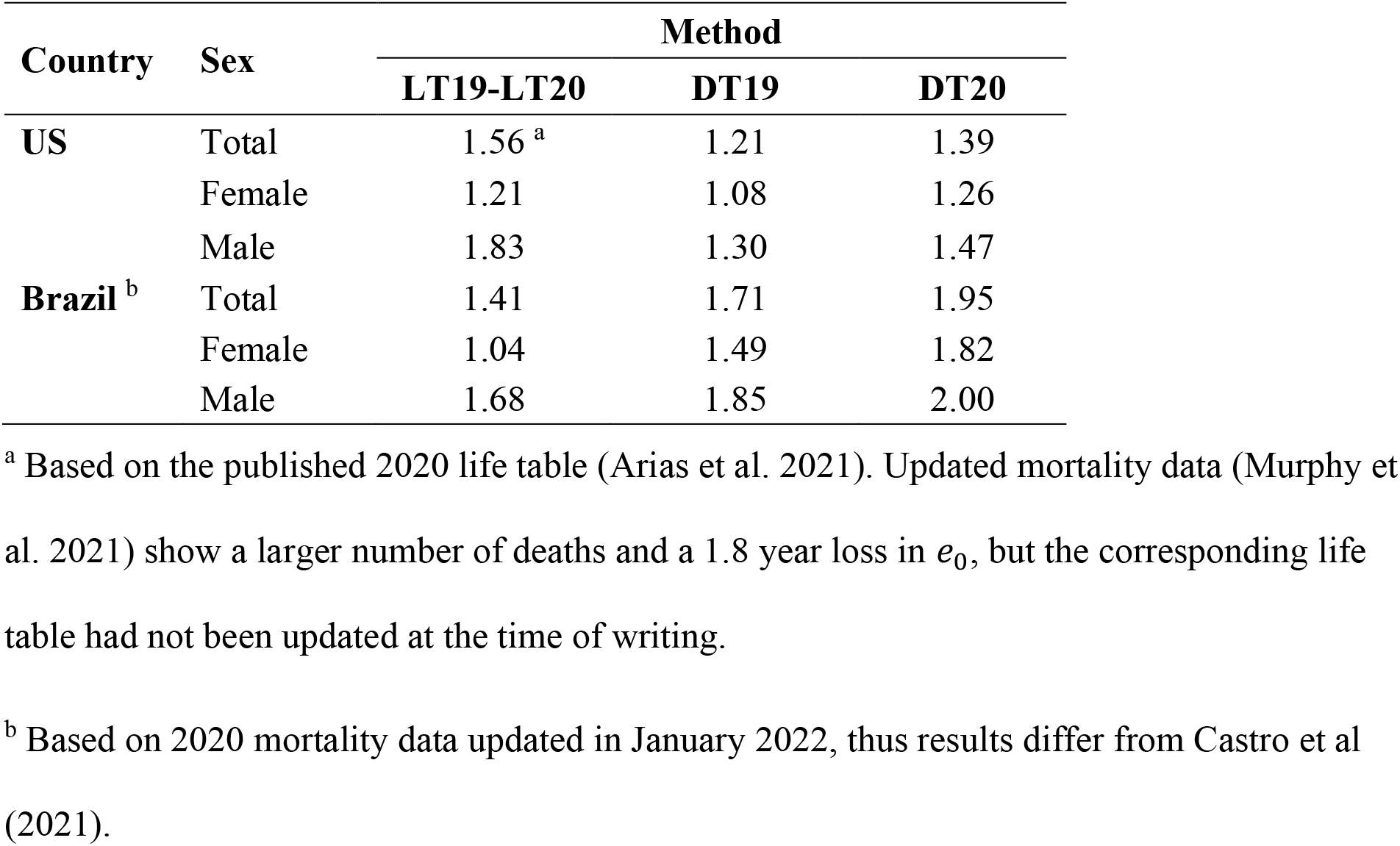
Estimated decline in *e*_0_ between 2019 and 2020 based on three methods, US and Brazil, by sex.

Between 2019 and 2020, age-specific mortality rates from all causes combined excluding COVID-19 increased for most age groups in the US, with substantial rises in the young adult and middle age groups (**Figure 1a**). In Brazil, however, rates increased only modestly in the young adult and middle age groups and declined at younger and older ages. The decline at younger ages in Brazil was mostly driven by a reduction in deaths due to influenza, pneumonia, and chronic and other lower respiratory diseases, likely a result of physical distancing and school closure.

**Fig. 1.**
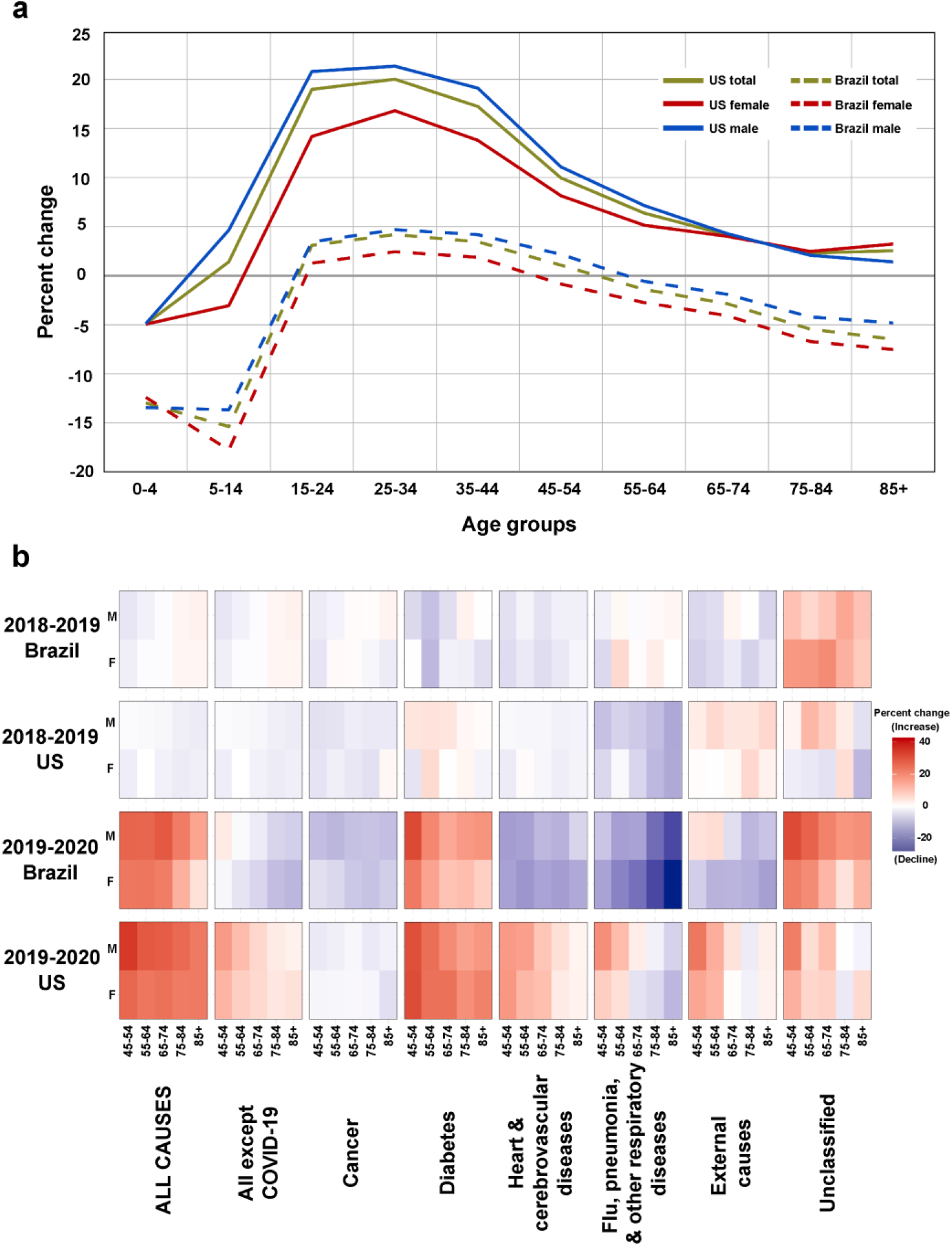
Percent change in age-specific death rates. **(a)** Between 2019 and 2020 for all non-COVID-19 causes combined, US and Brazil, by sex. **(b)** Heat map of the percent change in age-specific death rates between 2018-2019 and 2019-2020, for major categories of cause of death, US and Brazil, by sex, ages 45+.

Both the US and Brazil experienced large increases in diabetes mortality between 2019 and 2020, considerably more than between 2018 and 2019 (**Figure 1b**). Both countries, especially Brazil, experienced declines between 2019 and 2020 in cancer mortality. In contrast, Brazil generally faced declines in the following causes between 2019 and 2020 whereas the US generally saw increases: heart and cerebrovascular diseases; influenza, pneumonia, and other respiratory diseases; and external causes. Not all of these changes were necessarily a result of the pandemic (e.g., a potential secular decline in mortality), but most were likely due directly or indirectly to COVID-19.

Results show that DT19 and DT20 underestimated the decline in *e*_0_ for the US and overestimated the decline for Brazil relative to LT19-LT20. As shown in **Figure 1**, COVID-19 is not independent of other CoD. Since mortality rates from other causes generally increased between 2019 and 2020 in the US, the assumption of independence underestimates the overall change in mortality, whereas the opposite occurs in Brazil.

## Discussion

There are many potential reasons for the dependence between risks of dying from COVID-19 and those from other causes. Because COVID-19 fatality increases in the presence of co-morbidities (e.g., cancer, heart disease, diabetes), rates from these chronic ailments might have decreased if patients succumbed to COVID-19, a phenomenon related to “harvesting” (Schwartz 2000). Death rates from some external causes, such as violence and travel-related accidents, may have declined because of reduced social and work activities. In contrast, mortality rates from non-COVID-19 causes may have increased because of worsening of co-morbidities due to the effects of COVID-19; delays in primary/preventative care and reduced disease management for non-COVID-19 conditions; inadequate care in clinics and hospitals due to shortages of equipment, staff, and space; increased mortality from other causes due to long COVID-19; higher rates of smoking, drinking, drug use, and poor nutrition; reduced exercise; and potential health consequences from job loss, financial difficulties, and reduced social ties (Dey and Davidson 2021; Griffin 2021).

Our results are likely affected by the quality of reported death information. Misidentification or miscoding of CoD – particularly between COVID-19 and other respiratory conditions before widespread testing for the virus – could have inflated or reduced numbers of COVID-19 deaths. Unclassified deaths could have biased the estimates, although there has been little change in the relative size of this category between 2019 and 2020 (5.8% in Brazil for both years, and 1.1% and 1% in the US in 2019 and 2020, respectively). Underreporting of deaths, potentially selective by CoD and age, could have affected the age-specific pattern of both COVID-19 and non-COVID-19 mortality. Underreporting was estimated at 1.39% in 2019 for Brazil (IBGE 2019) and less than 1% in 2015-2018 for the US (Karlinsky 2021), but could have increased during the pandemic.

## Conclusions

Our results underscore that, despite the continued use of multiple decrement methods, they have serious drawbacks for determining the overall effect of the pandemic on *e*_0_. Especially troubling is that our findings suggest that the assumption of independence between COVID-19 and other causes could either underestimate or overestimate the overall loss in *e*_0_. Analyses that estimate declines in *e*_0_ due to COVID-19 when all CoD data are not available need to explicitly acknowledge this limitation.

## Data Availability

The data and code required to reproduce the results presented in this manuscript are available at 39 https://github.com/mcastrolab/Covid-19_competing_risk.

## Acknowledgment

TA acknowledges funding support from the National Institute on Aging under Award Number T32AG000037. The content is solely the responsibility of the authors and does not necessarily represent the official views of the National Institutes of Health.

